# Integrative analysis of circulating proteolytic biomarkers and genomic landscape in colorectal cancer

**DOI:** 10.64898/2026.07.14.26357715

**Authors:** E.D. Pankratova, K.A. Rubina, V.V. Kakotkin, M.A. Agapov, P.S. Klimovich, V.Yu. Sysoeva, A. Kashchenko, E.V. Semina

## Abstract

Colorectal cancer (CRC) is highly heterogeneous at both clinical and molecular levels, and the integration of circulating biomarkers with comprehensive genomic profiling remains limited. In this study, we measured circulating urokinase-type plasminogen activator (uPA) and its receptor (uPAR) in 53 patients with colorectal neoplasms and performed whole-genome sequencing (WGS) on matched tumor-normal pairs from 51 patients to characterize somatic mutations, copy number alterations (CNAs), tumor mutational burden (TMB), microsatellite instability (MSI), homologous recombination deficiency (HRD), and mutational signatures.

Circulating uPAR levels were significantly elevated in patients with CRC compared with healthy controls, showing a stepwise increase across tumor stages and reaching the highest levels in stage IV disease. In contrast, circulating uPA levels showed only a non-significant trend toward elevation and did not vary significantly by stage. Despite the strong association between uPAR and tumor progression, circulating uPA and uPAR levels were not significantly correlated with TMB, MSI, HRD scores, or the mutational status of major CRC driver genes, including TP53, KRAS, FBXW7, BRAF, NRAS, and PIK3CA. Genomic analysis revealed a heterogeneous mutational landscape dominated by TP53 and APC, with only a minority of tumors exhibiting high TMB or MSI. Mutational signatures were primarily clock-like (SBS1, SBS5), with minimal contribution from MMR- or HRD-related processes.

Together, these findings indicate that circulating uPAR is a robust marker of CRC progression that appears to operate largely independently of established genomic instability metrics. This supports uPAR potential utility in risk stratification and biological monitoring when integrated with molecular profiling.

## Introduction

Colorectal cancer (CRC) remains one of the leading malignancies of the digestive system, characterized by pronounced clinical and biological heterogeneity, as well as variable rates of disease progression (1). Although CRC incidence has generally declined among older populations in high-income countries, it remains the third most common cancer worldwide. In contrast, its incidence is increasing in countries with developing economies. Of particular concern is the rising number of cases among individuals under 50 years of age, underscoring the need for a deeper understanding of risk factors, including lifestyle changes, dietary patterns, and metabolic disturbances (2, 3).

The biological complexity of CRC is reflected in substantial variability in disease course and treatment response, highlighting the need for comprehensive characterization of molecular determinants underlying tumor behavior. This molecular heterogeneity is evident at both the genomic level and clinical phenotype of the disease. The most frequently observed somatic mutations occur in the APC, KRAS, TP53, and PIK3CA genes, which play critical roles in regulating cell proliferation, apoptosis, and key signaling pathways essential for maintaining intestinal epithelial homeostasis. The accumulation of alterations in these genes underlies the adenoma–carcinoma sequence and defines the principal pathways of colorectal tumorigenesis (4, 5).

Beyond canonical driver mutations, increasing attention has been directed toward the status of the DNA mismatch repair (MMR) system, particularly deficient mismatch repair (dMMR), and microsatellite instability-high (MSI-H) tumors. These molecular features, present in approximately 10–15% of CRC cases, are strongly associated with elevated mutational burden, neoantigen accumulation, and enhanced immune activation. Importantly, dMMR/MSI-H status serves as a robust predictive biomarker for response to immune checkpoint inhibitors (ICI) (6–8). However, a substantial proportion of dMMR/MSI-H tumors exhibit either primary or acquired resistance to immunotherapy, highlighting the complexity of underlying biological mechanisms and the need to identify additional predictive biomarkers and resistance pathways (9, 10). In addition to MSI/dMMR status, tumor mutational burden (TMB) represents an important component of molecular tumor profiling. TMB reflects the total number of somatic mutations within the tumor genome and is considered an integrative measure of genomic instability and tumor immunogenic potential. Elevated TMB is associated with increased neoantigen load and may enhance antitumor immune responses Nevertheless, the clinical relevance of TMB in CRC remains a subject of ongoing debate. High TMB is predominantly observed in MSI-H/dMMR tumors, whereas in microsatellite-stable (MSS) tumors, its association with prognosis and therapeutic response remains inconsistent (11, 12).

In addition to genetic determinants, components of extracellular matrix and tissue remodeling systems play a crucial role in tumor progression and aggressiveness. A prime example is the uPA/uPAR axis, comprising the urokinase-type plasminogen activator (uPA) and its receptor (uPAR, also known as CD87). Initially characterized as part of the fibrinolytic cascade, this system is now recognized as a key regulator of tumor invasion, migration, angiogenesis, and metastasis (13, 14). uPAR is a glycosylphosphatidylinositol (GPI)-anchored membrane protein that interacts with uPA, integrins, and other extracellular matrix components, thereby modulating both intracellular signaling pathways and extracellular matrix remodeling (15). Mounting evidence indicates that elevated levels of uPAR and its soluble form (suPAR) are associated with multiple malignancies and correlate with more aggressive tumor phenotypes and poorer clinical outcomes, including in CRC (16–18). Furthermore, molecular studies have demonstrated that *PLAUR* gene expression, which (encodes uPAR, is associated with clinicopathological features and survival outcomes in CRC patients. Functional studies further suggest that *PLAUR* suppression can inhibit tumor growth and invasiveness through modulation of key signaling pathways, including AKT/p53 signaling (19). Despite accumulating evidence, the role of circulating uPA and uPAR levels in biomonitoring CRC progression remains insufficiently studied, particularly in combination with TMB, MSI, and other molecular markers. Important questions remain as to whether changes in these biomarker levels reflect tumor aggressiveness regarding whether they are associated with the immune microenvironment, and whether they can serve as independent prognostic indicators for patient risk stratification.

Given the growing interest in integrating molecular and biochemical markers into clinical practice and personalized CRC treatment, it is important to comprehensively assess the associations between conventional genomic tumor characteristics, including TMB and MSI, proteolytic-system biomarkers such as uPA/uPAR, and clinical outcomes. Therefore, the aim of the present study was to assess circulating uPA and uPAR levels in patients with colorectal cancer, analyze their association with disease stage and molecular tumor characteristics, and determine their potential prognostic and clinical significance.

## Materials and Methods

### Study Design and Patient Cohort

This study had a cross-sectional design and was conducted in the Kaliningrad region within the framework of the “National Genetic Initiative” (NGI) population genomic program (20). Patients were recruited as part of the prospective observational study “Factors Affecting the Results of Treatment of Patients With Colorectal Cancer” (ClinicalTrials.gov identifier: NCT06050447). The study was conducted at Immanuel Kant Baltic Federal University in Kaliningrad in 2023. The study protocol was approved by the Local Ethics Committee of Immanuel Kant Baltic Federal University (Protocol No. 43, dated October 18, 2023). All participants provided written informed consent for the use of biological material and genetic data for research purposes.

Inclusion criteria comprised age ≥18 years, presence of an epithelial colorectal neoplasm detected by colonoscopy (types 2B–3 according to the JNET classification and pit pattern Vi or Vn according to the S. Kudo classification), and subsequent histopathological verification of colorectal adenocarcinoma or adenoma. Non-inclusion criteria included age <18 years, presence of parenterally transmitted infections (HIV infection, hepatitis B or C, or syphilis), refusal to participate, inability to provide informed consent due to cognitive or other limitations, and insufficient biological material for morphological assessment and DNA extraction. Exclusion criteria included the absence of tumor growth in biopsy specimens, as determined by pathological examination, and withdrawal of consent for the use of sequencing data at any stage of the study. Patients were recruited at the stage of preparation for endoscopic examination. Following informed consent and identification of a neoplasm, biological samples were collected. For each patient, paired samples were obtained: a tumor biopsy specimen (∼2–3 mm³) from the most affected mucosal region of the colon and a peripheral venous blood sample (16 mL), which served as a matched normal control for germline variant analysis.

A control cohort consisting of 16 apparently healthy volunteers was included for comparative analysis of circulating uPA and uPAR levels. Control participants had no known history of malignant disease, inflammatory bowel disease, or other active inflammatory conditions at the time of blood collection. Peripheral venous blood samples were collected after written informed consent and processed using the same protocols as those applied to the colorectal cancer patient cohort. Healthy controls were included exclusively for the assessment of circulating urokinase system biomarkers and were not subjected to genomic analyses.

### Nucleic Acid Extraction

Genomic DNA was extracted using a magnetic bead–based method with the MGIEasy Magnetic Beads Blood Genomic DNA Extraction Kit (MGI). DNA quality and concentration were assessed by spectrophotometry and fluorometry prior to library preparation. Whole-genome sequencing libraries were prepared using the MGIEasy FS PCR-Free Library Prep Set (96 reactions, MGI). Sequencing was performed on the DNBSEQ-T7 platform in paired-end mode (2 × 150 bp).

### Whole-genome sequencing and data processing

Raw reads were preprocessed using fastp (v0.23.2) (https://github.com/OpenGene/fastp) (21). Adapter trimming and quality filtering were performed using the parameters “-5 -3 -W 4 -M 20 - l 70 -q 20 -u 20”. Reads were aligned to the human reference genome GRCh38.p14 using BWA-MEM (v0.7.17) (https://github.com/lh3/bwa) with parameters “-M -v 3” (22). Alignment files were converted to BAM format, sorted, and indexed using samtools (v1.16) (https://github.com/samtools/samtools) (23). PCR duplicates were identified and marked using MarkDuplicates from the Picard toolkit (v2.27) (https://broadinstitute.github.io/picard/) with the parameter “-MAX_RECORDS_IN_RAM 50000000”. Sequencing data quality was assessed at multiple stages using FastQC (v0.11.9) (https://www.bioinformatics.babraham.ac.uk/projects/fastqc/) and FastQ Screen (v0.14.0) (https://github.com/StevenWingett/FastQ-Screen) (24), while genome coverage was calculated using Mosdepth (v0.3.3) (Pedersen and Quinlan, 2018; https://github.com/brentp/mosdepth) (25).

### Somatic Variant Calling

Germline single-nucleotide variants (SNVs) and short insertions/deletions (indels) were identified using DeepVariant (v1.4) (https://github.com/google/deepvariant) in whole-genome sequencing mode (--model_type=WGS) (26). Somatic SNVs and indels were detected using Strelka2 (v2.9.10) (https://github.com/Illumina/strelka) in paired tumor–normal mode with default parameters (27). A variant allele frequency (VAF) threshold of 5% was applied, as mutation detection becomes less reliable below this level, particularly at a sequencing depth of approximately 100×. Functional annotation of genetic variants was performed using the Ensembl Variant Effect Predictor (VEP, release 110) (https://github.com/Ensembl/ensembl-vep) with the following parameters: --sift b --polyphen b --protein --symbol --ccds --uniprot --per_gene --domains --check_existing --clin_sig_allele 1 --max_af --af_1kg --af_gnomade --pubmed --var_synonyms --failed 1 (28). Clinical interpretation of genomic alterations was performed using the CIViC (Clinical Interpretation of Variants in Cancer) database (https://civicdb.org) and OncoVar (https://oncovar.org). Somatic variants were additionally annotated using the COSMIC database (Catalogue of Somatic Mutations in Cancer, version 103) and the Cancer Gene Census (CGC), which contains genes with established roles in carcinogenesis (https://cancer.sanger.ac.uk/cosmic). Only coding somatic variants identified in genes annotated in COSMIC and included in the Cancer Gene Census were retained for downstream analyses.

### Copy Number Alteration Analysis

Copy number alterations (CNAs) were identified using CNVkit (v0.9.10) (https://github.com/etal/cnvkit) (29). The analysis was performed in batch mode (cnvkit.py batch) with parameters “-m wgs --annotate refflat38.txt --target-avg-size 1000”. Annotation of CNVs and estimation of gene copy numbers were based on the GRCh38 genome annotation obtained from the UCSC Genome Browser (https://hgdownload.soe.ucsc.edu/downloads.htm).

### Microsatellite Instability (MSI) Analysis

MSI status was assessed using MSIsensor-pro (v1.2) (https://github.com/xjtu-omics/msisensor-pro) (30). Microsatellite loci were first identified across the GRCh38.p14 reference genome using the scan function. Tumor–normal paired analysis was then performed using the msi function with default parameters. For each sample, the total number of microsatellite loci, the number of unstable loci, and the proportion of unstable sites (MSIsensor score) were calculated. Tumors with an MSIsensor score below 3.5 were classified as microsatellite stable (MSS), whereas samples with a score ≥3.5 were considered MSI-positive.

### Homologous Recombination Deficiency (HRD) Analysis

Homologous recombination deficiency was assessed using HRDCNA (https://github.com/XSLiuLab/HRDCNA), which estimates the probability of HR pathway disruption based on genome-wide copy number profiles (31). Segmented copy number data (*.call.cns) generated by CNVkit were used as input. The algorithm outputs an HRDCNA score ranging from 0 to 1, with higher values indicating a greater likelihood of homologous recombination deficiency.

### Tumor mutational burden (TMB)

Tumor mutational burden (TMB) was defined as the number of somatic mutations per megabase (Mb) of the analyzed genomic sequence (32). Only somatic variants identified in tumor samples were included, following exclusion of germline variants using matched normal blood samples and standard quality filtering criteria. Samples were stratified into TMB categories using the following thresholds: low (<10 mutations/Mb), intermediate (10–20 mutations/Mb), high (20–30 mutations/Mb), very high (30–100 mutations/Mb), and ultra-high (≥100 mutations/Mb).

### Mutational signature analysis

Mutational signatures were analyzed using SigProfilerAssignment (v0.0.29) (https://github.com/AlexandrovLab/SigProfilerAssignment) (33). This analysis was performed on somatic variants in VCF format using the COSMIC mutational signatures database v3.4 (https://cancer.sanger.ac.uk/signatures/sbs/), allowing the contribution of individual mutational processes to be quantified for each sample.

### Determination of serum uPA and uPAR levels

Serum concentrations of urokinase-type plasminogen activator (uPA) and urokinase plasminogen activator receptor (uPAR) were determined by enzyme-linked immunosorbent assay (ELISA) using commercial kits (ab108917 and ab246549, Abcam, USA) according to the manufacturer’s protocols. Serum was obtained from peripheral blood samples by clotting and centrifugation and stored at −80°C until analysis. Absorbance was measured at 450 nm using a Bio-Rad microplate reader (USA), and analyte concentrations were calculated from standard curves. All measurements were performed in duplicate.

### Statistical analysis

Statistical analysis was performed using GraphPad Prism software (version 9.0, GraphPad Software, USA). The normality of the distribution of quantitative variables was assessed using the Shapiro–Wilk test. Since the distribution of both uPA and uPAR concentrations in the studied groups significantly deviated from normal, nonparametric statistical methods were used for further analysis. The Mann–Whitney U test was employed to compare two independent groups (patients with oncological diseases and healthy volunteers). Results were presented as medians and interquartile ranges [Q1; Q3]. Differences were considered statistically significant at p < 0.05.

## Results

### Patient Cohort and Clinicopathological Characteristics

A total of 53 patients with epithelial colorectal neoplasms detected during colonoscopy were initially enrolled in the study. Circulating levels of uPA and uPAR were measured in all patients. However, whole-genome sequencing data of sufficient quality for downstream genomic analyses could not be obtained from two tumor tissue samples. Consequently, analyses of clinicopathological characteristics, somatic mutations, tumor mutational burden (TMB), microsatellite instability (MSI), homologous recombination deficiency (HRD), mutational signatures, and copy number alterations (CNAs) were performed for 51 patients, whereas analyses of circulating uPA and uPAR levels included the entire cohort of 53 patients.

The median age of 71 years. The study cohort comprised 24 women and 27 men. Individual clinicopathological characteristics of the 51 patients included in the genomic analyses are provided in the Supplementary Table S1. A summary of the clinicopathological characteristics of patients with colorectal cancer is presented in Table 1.

### Clinical and pathological characteristics of patients with colorectal cancer (CRC)

| Characteristic | Category | n (%) |
| --- | --- | --- |
| Tumor localization | Rectum | 12 (23.53%) |
|  | Rectosigmoid junction | 6 (11.76%) |
|  | Sigmoid colon | 13 (25.49%) |
|  | Upper colon | 20 (39.22%) |
| Tumor grade | High grade | 5 (9.8%) |
|  | Low grade | 46 (90.2%) |
| Tumor stage | I | 3 (5.88%) |
|  | II | 15 (29.41%) |
|  | III | 18 (35.29%) |
|  | IV | 15 (29.41%) |
| pT stage | pT2 | 8 (15.69%) |
|  | pT3 | 29 (56.86%) |
|  | pT4a | 11 (21.57%) |
|  | pT4b | 3 (5.88%) |
| pN stage | pN0 | 21 (41.18%) |
|  | pN1a | 15 (29.41%) |
|  | pN1b | 4 (7.84%) |
|  | pN1c | 2 (3.92%) |
|  | pN2a | 4 (7.84%) |
|  | pN2b | 5 (9.8%) |
| Metastasis (M stage) | pM0 | 36 (70.59%) |
|  | pM1a | 7 (13.73%) |
|  | pM1b | 7 (13.73%) |
|  | pM1c | 1 (1.96%) |
| Lymphovascular invasion | L0 | 31 (60.78%) |
|  | L1 | 20 (39.22%) |
| Vascular invasion | V0 | 45 (88.24%) |
|  | V1 | 6 (11.76%) |
| Perineural invasion | Pn0 | 43 (84.31%) |
|  | Pn1 | 8 (15.69%) |
| Tumor budding | Bd1 | 24 (47.06%) |
|  | Bd2 | 17 (33.33%) |
|  | Bd3 | 10 (19.61%) |

### Sequencing quality and data processing

Whole-genome sequencing achieved a mean coverage depth of 100× for tumor tissue samples and 30× for matched peripheral blood samples.

### Somatic Mutation Landscape

After applying the filtering criteria (variant allele frequency >5% and at least five supporting reads), a total of 17,318 somatic mutations were identified across the cohort. The mean number of mutations per patient was 339.57, whereas the median number was 161 (Supplementary Table S1).

Analysis of mutation frequencies across individual genes revealed that TP53 and APC were the most frequently mutated genes in the cohort, occurring in 84.3% and 64.7% of patients, respectively. Mutations in TTN (47.1%) and KRAS (41.2%) were also common. In addition, mutations in MUC4 were detected in 39.2% of patients, while MUC16, MUC19, and MUC6 were each mutated in 29.4% of cases. Several other genes, including FAT4, SYNE1, and OBSCN, harbored mutations in approximately 20–22% of patients (Supplementary Table S3).

Analysis of recurrent variants demonstrated that KRAS mutations were the most frequent individual alterations in the cohort. The KRAS variant NC_000012.12:g.25245350C>A (NM_004985.5:c.35G>T, p.Gly12Val) was identified in 8 patients (15.7%), whereas the NC_000012.12:g.25245350C>T (NM_004985.5:c.35G>A, p.Gly12Asp) variant was detected in 3 patients (5.9%). Recurrent mutations were also observed in TP53 (NC_000017.11:g.7673802C>T, NM_000546.6:c.818G>A, p.Arg273His), which was present in 5 patients (9.8%), as well as in BRAF (NC_000007.14:g.140753336A>T, NM_004333.6:c.1799T>A, p.Val600Glu), FBXW7 (NC_000004.12:g.152328232C>T, NM_001349798.2:c.1394G>A, p.Arg465His), and APC (NC_000005.10:g.112792446C>T, NM_000038.6:c.646C>T, p.Arg216* and NC_000005.10:g.112839942C>T, NM_000038.6:c.4348C>T, p.Arg1450*), which were identified in 4 (7.8%), 3 (5.9%), and 3 (5.9%) patients, respectively.

Additional recurrent variants were detected in several genes, including KLF18, NBPF3, CACNA1C, CARMIL1, FNIP1, PPP1R12B, and ZNF518A. A complete list of recurrent somatic variants ranked by decreasing frequency is provided in Supplementary Table S4.

### Annotation of Somatically Altered Genes Using COSMIC and IntOGen

To further characterize the biological significance of the identified somatic alterations, genes were annotated using the COSMIC Cancer Gene Census (CGC) and the IntOGen platform (Supplementary Tables S5–S6). This analysis aimed to identify genes with established or putative roles in tumorigenesis.

According to the COSMIC Cancer Gene Census, 435 genes harboring somatic variants in the study cohort were annotated as oncogenes, tumor suppressor genes (TSGs), and/or genes involved in gene fusion events. Analysis using the IntOGen database revealed that 412 mutated genes were classified as driver genes, indicating evidence of positive selection during tumor development and a potential contribution to tumorigenesis.

Overall, integration of COSMIC and IntOGen annotations identified 114 genes previously associated with colorectal cancer, highlighting the involvement of both well-characterized and potentially novel molecular mechanisms in the pathogenesis of the analyzed tumors. Of these, 37 genes were additionally classified as driver genes.

### Driver and clinically relevant mutations

Analysis of driver mutations based on OncoVar annotations identified 49 driver genes. Somatic mutations affecting at least one of these genes were detected in 44 of 51 patients (86.3%). The most frequently altered driver genes were TP53 and KRAS, which were mutated in 29 (56.9%) and 19 (37.2%) patients, respectively. Less frequent alterations were observed in FBXW7, which was mutated in 6 patients (11.8%), as well as in BRAF and NRAS, each affected in 4 patients (7.8%) (Supplementary Tables S1–S2).

According to the CIViC database, clinically relevant biomarker mutations were identified in 33 of 51 patients (64.7%). The most common alterations were KRAS G12V mutations, detected in 8 patients (15.7%), and TP53 R273H mutations, identified in 5 patients (9.8%). Recurrent alterations also included BRAF V600E (7.8%), as well as KRAS G12D, KRAS G13D, TP53 G245S, and TP53 R175H, each observed in 5.9% of cases. A complete list of clinically relevant biomarker mutations ranked by decreasing frequency is provided in Supplementary Table S2.

### Copy Number Alterations

On average, 3,045 gene amplifications and 3,053 gene deletions were identified per patient. Among genes annotated in COSMIC as oncogenes, the most frequently amplified were GNAS (70.59%), SALL4 (70.59%), SRC (70.59%), NFATC2 (70.59%), CARD11 (47.06%), ETV1 (47.06%), EGFR (47.06%), PREX2 (43.14%), UBR5 (41.18%), and BRAF (39.22%). Less frequent amplifications affected SETDB1 (15.69%), BCL9 (13.73%), ERBB2 (11.76%), NTRK2 (7.84%), RSPO3 (5.88%), PIK3CA (7.84%), KRAS (7.84%), and PTPN11 (3.92%).

Alterations involving EGFR, ERBB2, SRC, and NTRK2 may reflect activation of receptor tyrosine kinase signaling associated with the activation of the MAPK/ERK and PI3K/AKT cascades (34–39). Amplifications of KRAS, BRAF, and PIK3CA may further enhance signaling through downstream components of the same pathways, specifically the RAS–RAF–MEK–ERK axis for KRAS/BRAF and the PI3K/AKT pathway for PIK3CA (40, 41). GNAS and PREX2 are additionally involved in the regulation of secondary messenger systems and PI3K-dependent signaling, whereas SALL4 and SETDB1 are associated with transcriptional and epigenetic regulation and may contribute to maintaining a proliferative and less differentiated state of tumor cells (42–46).

Amplifications of genes annotated as tumor suppressor genes (TSGs) were also common and included ASXL1 (70.59%), PTPRT (70.59%), CSMD3 (41.18%), KMT2C (37.25%), AXIN1 (15.69%), GRIN2A (15.69%), RNF43 (9.8%), TSC1 (7.84%), APC (3.92%), SMAD2 (3.92%), TGFBR2 (3.92%), PIK3R1 (3.92%), POLD1 (3.92%), CDKN2A (3.92%), ATM (1.96%), and several others.

The presence of amplifications in tumor suppressor genes should be interpreted with caution, as such alterations do not necessarily imply increased gene function. Rather, they most reflect large-scale chromosomal rearrangements or co-amplification of the neighboring genomic regions. Nevertheless, recurrent alterations involving APC, AXIN1, and RNF43 suggest potential involvement of the WNT/β-catenin pathway, whereas amplifications of TGFBR2 and SMAD2 indicate perturbations in TGF-β signaling, which plays a complex and context-dependent role in colorectal cancer (47–53).

A separate category consisted of genes with dual oncogenic and tumor-suppressive functions, including RAD21 (41.18%), CUX1 (39.22%), NOTCH1 (9.8%), MAP3K1 (3.92%), TBX3 (3.92%), and QKI (5.88%). These genes are involved in the regulation of transcription, cell-cycle progression, and signal transduction pathways, and their amplification may lead either to activation or disruption of regulatory mechanisms depending on the cellular context (54–65).

Among deletions, tumor suppressor genes (TSGs) were most frequently affected, including AMER1 (56.86%), SMAD4 (56.86%), SMAD2 (54.90%), RBM10 (52.94%), BCOR (52.94%), ARHGEF10 (49.02%), LEPROTL1 (45.10%), and TP53 (39.22%). Additional genes affected by recurrent deletions included FBXW7 (21.57%), FAT4 (19.61%), APC (15.69%), ARID1A (13.73%), PIK3R1 (11.76%), CDKN2A (11.76%), ATM (11.76%), NCOR2 (5.88%), TGFBR2 (3.92%), POLD1 (3.92%), and ACVR2A (3.92%).

This deletion profile is consistent with classical mechanisms of tumor suppressor gene inactivation in cancer. In particular, frequent losses of TP53, ATM, and POLD1 suggest disruption of DNA damage response pathways and impaired mechanisms responsible for maintaining genome stability. Deletions affecting SMAD4 and SMAD2 indicate dysregulation of the TGF-β signaling pathway, which normally restricts the proliferation of intestinal epithelial cells. Losses involving APC, AMER1, and other components of the pathway are indicative of WNT/β-catenin pathway dysregulation, a key molecular event in colorectal tumorigenesis (50, 53, 66, 67).

Deletions affecting genes annotated as oncogenes were less common and included AR (56.86%), USP6 (35.29%), HIF1A (19.61%), AKT1 (19.61%), MTOR (13.73%), NRAS (7.84%), ERBB2 (7.84%), and PTPN11 (5.88%). These alterations most likely reflect nonspecific segmental losses arising as part of large chromosomal rearrangements rather than selective pressure for oncogene inactivation (47, 68).

Genes with mixed oncogenic and tumor-suppressive functions included MAP3K1 (11.76%), NOTCH1 (5.88%), TET1 (5.88%), CTNNB1 (3.92%), and TBX3 (3.92%).

A complete list of gene amplifications and deletions ranked by decreasing frequency is provided in Supplementary Tables S3–S4.

### TMB, MSI, and HRD Profiles

Analysis of tumor mutational burden (TMB), calculated as the number of coding somatic mutations per megabase of coding sequence, revealed a mean value of 14.97 mut/Mb (SD = 26.5, coefficient of variation = 177.02%), whereas the median value was substantially lower at 6.03 mutations/Mb (Q1–Q3: 4.5–8.46), indicating a markedly skewed distribution driven by a small subset of hypermutated tumors. Most tumors were classified as having low or intermediate mutational burden, with 18 of 51 cases (35.3%) categorized as Low and 24 cases (47.1%) as Intermediate. According to the FDA criterion for TMB-high status (≥10 mutations/Mb), 9 patients (17.65%) were classified as having a high mutational burden. FDA has approved pembrolizumab for adults and children with TMB-H solid tumors. Food and Drug Administration. 2020. Available at: https://www.fda.gov/drugs/drug-approvals-and-databases/fda-approves-pembrolizumab-adults-and-children-tmb-h-solid-tumors. The maximum TMB value reached 134.1 mutations/Mb (Supplementary Table S2).

The mean MSI score was 0.91 (SD = 2.1, coefficient of variation = 231.47%), with a median value of 0.07 (Q1–Q3: 0.0–0.52). Tumors with an MSIsensor score below 3.5 were classified as microsatellite stable (MSS), whereas samples with a score ≥3.5 were considered MSI-positive. The majority of tumors in the cohort were classified as MSS, consistent with the low median MSI score. Mutations in MSH6, POLG, PARP4, APC, and SMAD2 were identified in MSI-positive tumors. Among these, alterations in DNA repair and genome maintenance genes, particularly MSH6, represent the most biologically plausible explanation for the elevated MSI phenotype.

The mean HRDCNA score was 0.0021 (SD = 0.0031, coefficient of variation = 152.98%), with a median value of 0.0014 (Q1–Q3: 0.0014–0.0014). The maximum HRDCNA score was 0.0234. Overall, HRDCNA scores remained low across the cohort.

These findings point to a substantial heterogeneity in tumor mutational burden and genomic instability within the study cohort. Despite the relatively high mean TMB value, the distribution was markedly skewed by a limited number of hypermutated tumors, whereas most cases exhibited low or intermediate mutational burden (Supplementary Table S2).

MSI scores were generally low, indicating a predominance of microsatellite-stable tumors (MSS) and the absence of widespread mismatch repair (MMR) deficiency in the majority of cases.

Similarly, HRDCNA scores remained low, suggesting a lack of significant defects in homologous recombination repair and indicating a limited contribution of HRD-associated mechanisms to genomic instability in this cohort.

Taken together, these findings suggest that the cohort was predominantly composed of tumors with moderate mutational burden and without pronounced MSI- or HRD-associated genomic instability, whereas hypermutated and genomically unstable tumor subtypes represented a minority of cases.

To further characterize the mutational landscape across the cohort, we constructed a gene-level mutation heatmap of recurrently altered genes (Fig. 1). The heatmap illustrates the distribution of somatic mutations across individual patients, along with key clinicopathological and molecular features, including age, sex, tumor stage, grade, and TMB category.

**Figure 1.**
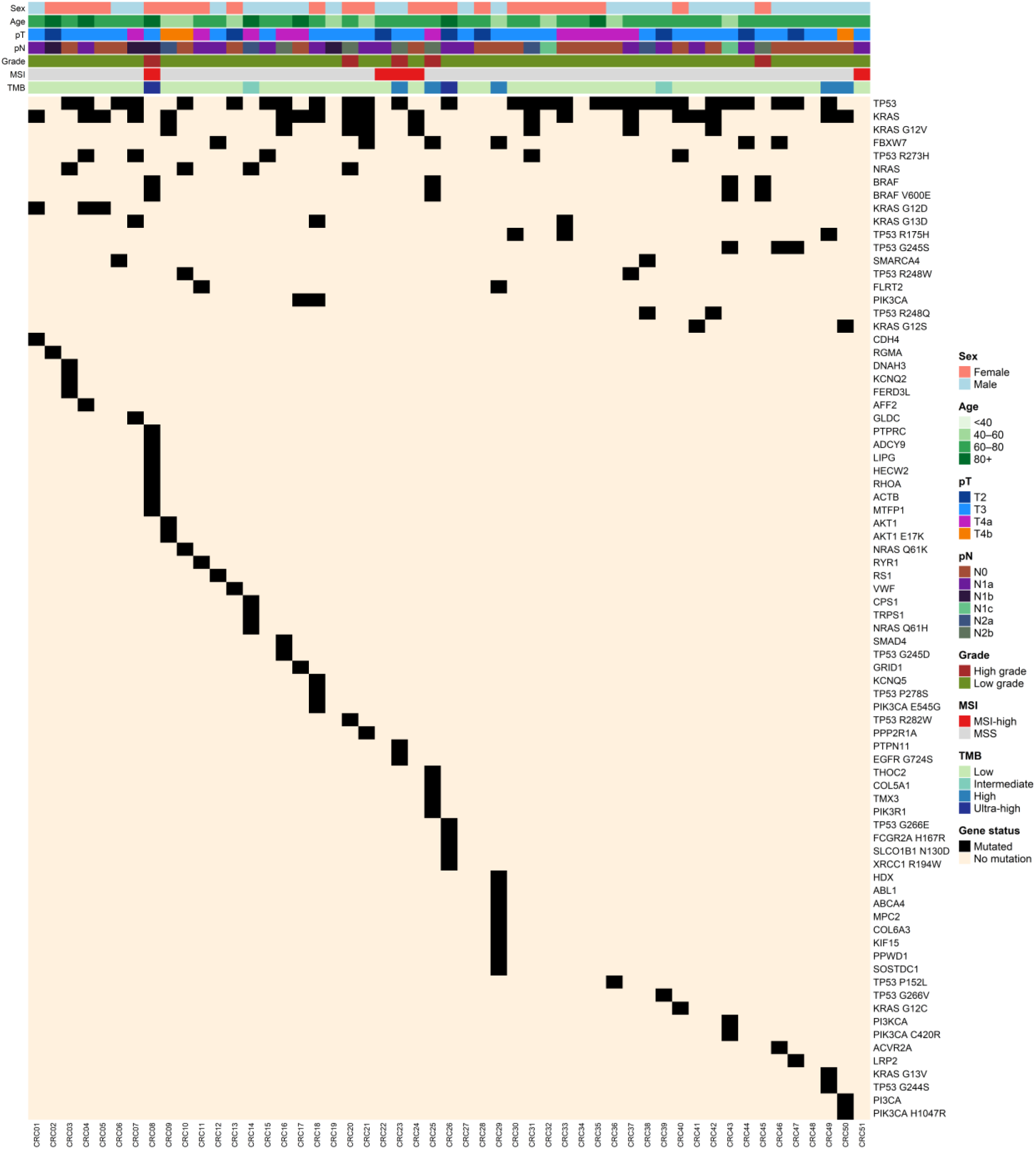
Mutational landscape of colorectal cancer cohort. Oncoplot showing the distribution of somatic mutations across individual patients. Each column represents an individual patient, and each row corresponds to a gene or recurrent variant. Black cells indicate mutated genes or variants, whereas beige cells indicate no detected mutation. Top annotations include clinical and molecular features: sex, age, pT stage, pN stage, tumor grade, and TMB category. Genes are ordered by decreasing mutation frequency. The plot highlights recurrent alterations in key colorectal cancer driver genes, including TP53, KRAS, and APC, as well as the overall heterogeneity of the mutational profiles across the cohort.

Recurrent mutations were most frequently observed in canonical CRC driver genes such as TP53, KRAS, and APC, with substantial inter-patient heterogeneity. No clear clustering of mutation patterns was observed based on TMB category or clinicopathological features, suggesting a high degree of genomic diversity within the cohort.

### Mutational Signatures

Analysis of mutational signatures revealed that the mutational landscape of the study cohort was predominantly shaped by endogenous mutagenic processes (Supplementary Tables S5–S6). The most frequently detected signatures were SBS1 and SBS5, which were present in 94.1% and 92.2% of patients, respectively. These findings are consistent with their established roles as clock-like mutational signatures reflecting the accumulation of somatic mutations associated with aging and cellular proliferative activity (69, 70).

The average contribution of SBS5 (0.32) and SBS1 (0.047) further supports the substantial role of baseline replicative processes in shaping the mutational profiles of these tumors.

SBS39 was detected in 44 of 51 patients (86.3%) and exhibited a high mean contribution (0.71). SBS40c was identified in only one patient (1.96%); however, its contribution within that sample reached 0.34. Previous studies have shown that SBS40 occurs across multiple tumor types, although its etiology remains unclear due to its similarity to SBS5 (70).

Mutational signatures associated with DNA repair defects were detected considerably less frequently. In particular, SBS3 (https://cancer.sanger.ac.uk/signatures/sbs/sbs3/), which is characteristic of homologous recombination deficiency, was identified in only 1.96% of patients, consistent with the low HRDCNA scores observed in the cohort (70).

Similarly, SBS6 (https://cancer.sanger.ac.uk/signatures/sbs/sbs6/) and SBS26 (https://cancer.sanger.ac.uk/signatures/sbs/sbs26/), which are associated with mismatch repair (MMR) deficiency and microsatellite instability, were detected only sporadically, in agreement with the low MSI scores observed in the study cohort (70).

Additional signatures, including SBS8, SBS33, SBS43, SBS57, SBS84, and SBS96, were also identified; however, both their frequency and contribution were low, suggesting a limited role for the corresponding mutational processes in the analyzed tumors.

Overall, these findings indicate that the mutational profiles of tumors in this cohort were predominantly driven by endogenous age-related mutational processes, whereas the contribution of specific DNA repair defects, including MMR deficiency and homologous recombination deficiency, appeared to be minimal. These observations were consistent with the MSI and HRDCNA results described above.

### Associations of serum uPAR/uPA levels with clinicopathological and genomic characteristics of colorectal cancer

Circulating levels of the urokinase system components uPA and uPAR were compared between healthy controls and patients with colorectal cancer (CRC). uPAR levels were significantly elevated in CRC patients compared with healthy individuals (mean ± SEM: 6117 ± 317 pg/mL, n = 53 vs 4292 ± 306 pg/mL, n = 16; P = 1.37 × 10⁻⁴, Fig. 2 A). In contrast, circulating uPA levels showed only a non-significant trend toward higher values in CRC patients (4.07 ± 0.67 ng/mL vs 2.42 ± 0.55 ng/mL, P = 0.062, Fig. 2 B). When stratified by pathologic stage, circulating uPAR levels increased with disease progression (Fig. 2 C). Notably, stage IV patients exhibited significantly higher uPAR levels compared with healthy controls (6758 ± 473 pg/mL vs 4292 ± 306 pg/mL, P = 0.013). In contrast, circulating uPA levels did not vary significantly across CRC stages (P = 0.536, Fig. 2 D). Together, these findings indicate that circulating uPAR, but not uPA, is significantly elevated in CRC patients and is associated with tumor progression.

**Figure 2.**
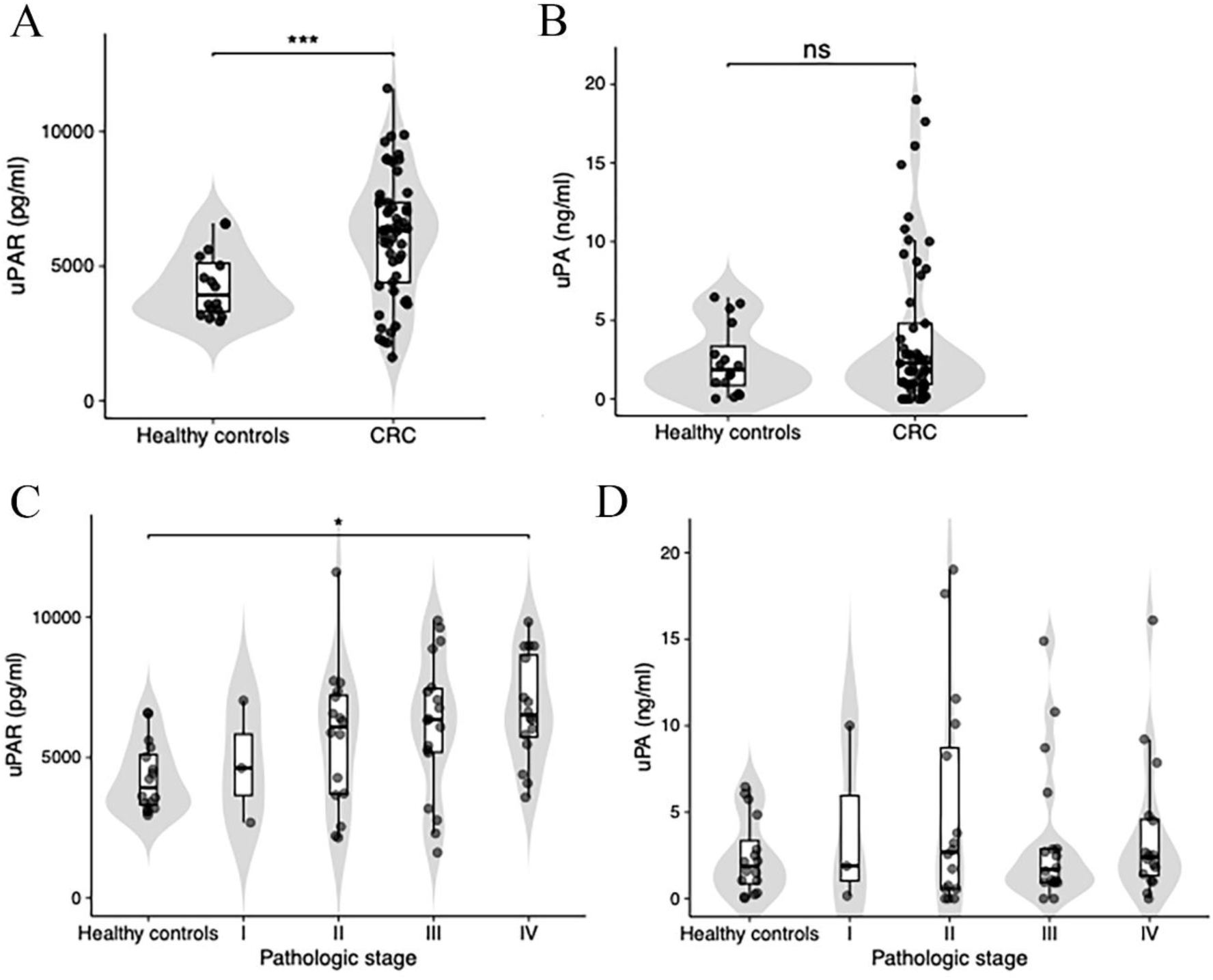
Circulating uPAR and uPA levels in healthy controls and colorectal cancer (CRC) patients. **A**, Circulating uPAR levels in healthy controls and CRC patients. **B**, Circulating uPA levels in healthy controls and CRC patients. **C**, Circulating uPAR levels by pathologic stage. **D**, Circulating uPA levels by pathologic stage. Each dot represents an individual sample. Box plots indicate the median and interquartile range, with whiskers extending to 1.5× the interquartile range. P values shown on the plots were calculated using t-test or one-way ANOVA, Tukey’s post hoc test. (ns, non-significant; *, p < 0.05; ***, p < 0.001)

We next assessed whether circulating uPAR and uPA levels were associated with genomic characteristics of the tumors. No significant correlations were observed between circulating uPA or uPAR levels and TMB, MSI, or HRD CNA score (Fig 3). Comparison of serum biomarker levels between tumors with wild-type and mutant status for recurrent CRC driver genes, including TP53, KRAS, FBXW7, BRAF, NRAS, and PIK3CA, revealed no significant differences in either uPAR or uPA levels (t-test, P > 0.05).

**Figure 3.**
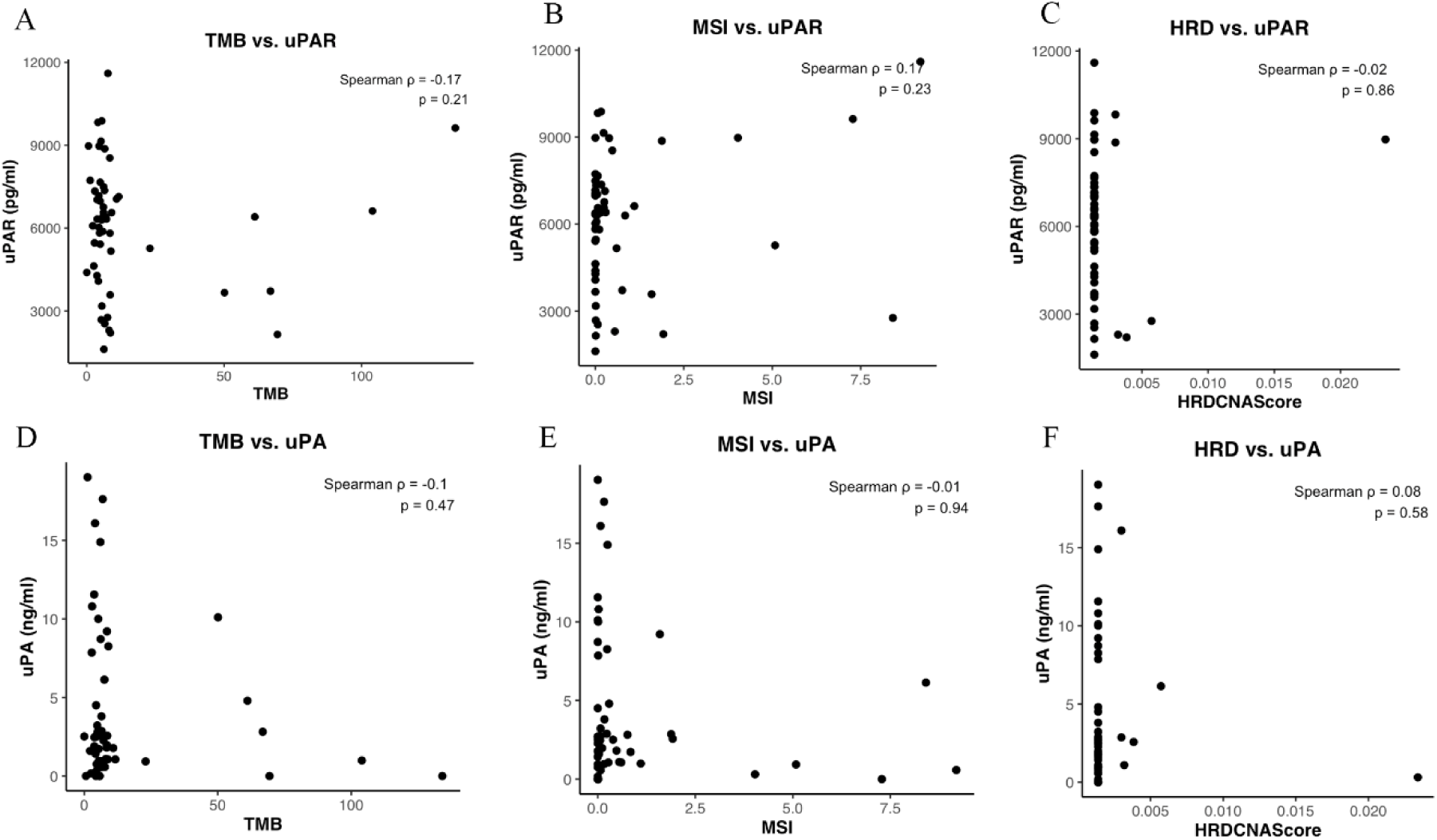
Correlation analysis between circulating uPA/uPAR levels and genomic features in colorectal cancer. Scatter plots showing the relationship between circulating uPAR **(A-C)** or uPA **(D-F)** levels and genomic variables including tumor mutational burden (TMB, **A, D**), microsatellite instability (MSI, **B, E**), and HRD CNA score (**C, F**). Each dot represents an individual sample. Associations were assessed using two-sided Spearman rank correlation, and the corresponding Spearman correlation coefficient (ρ) and p value are indicated.

We next examined whether mutations in recurrent colorectal cancer driver genes were associated with clinicopathologic tumor characteristics. Mutation frequencies for TP53, KRAS, FBXW7, BRAF, NRAS, and PIK3CA were compared across tumor features including age group, pathologic stage, metastatic status, lymphatic invasion, vascular invasion, perineural invasion, and tumor budding using Fisher’s exact tests (Fig 4). Overall, no statistically significant associations between driver gene mutations and tumor characteristics were detected after multiple testing correction (Benjamini-Hochberg). A nominal enrichment of BRAF mutations was observed in tumors with perineural invasion (Pn1) compared with Pn0 tumors (P = 0.0088); however, this association did not remain significant after correction for multiple comparisons (Padj = 0.37).

**Figure 4.**
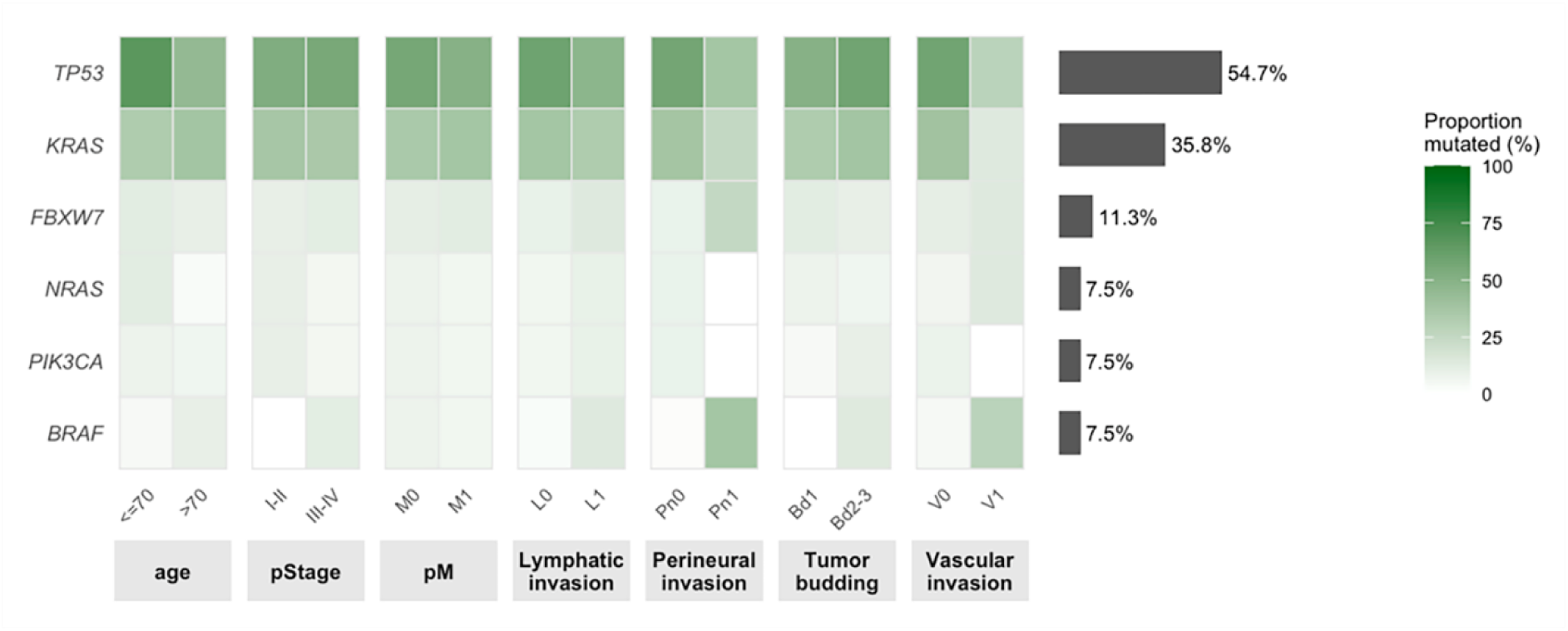
Distribution of CRC driver gene mutations across clinicopathologic tumor characteristics. Heatmap showing the proportion of mutated tumors (%) for recurrent CRC driver genes (BRAF, PIK3CA, NRAS, FBXW7, KRAS, TP53) across clinicopathologic features including age group, pathologic stage, metastatic status, lymphatic invasion, vascular invasion, perineural invasion, and tumor budding. Color intensity represents the percentage of tumors harboring a mutation in the indicated gene within each subgroup. The bar plot on the right summarizes the overall mutation prevalence for each gene. Associations between mutation status and tumor features were evaluated using Fisher’s exact tests with Benjamini-Hochberg correction. ** Nominal association between BRAF mutation and perineural invasion (P = 0.0088), not significant after multiple-testing correction (Padj = 0.37).

## Discussion

In this study, we performed a comprehensive molecular and biochemical characterization of colorectal cancer (CRC) by integrating genomic profiling with circulating biomarkers of the urokinase system. Our findings demonstrate that circulating uPAR levels are significantly elevated in CRC patients and are positively associated with tumor stage, supporting its potential role for uPAR as a marker of disease progression.

Notably, uPAR levels increased with advancing pathological stage, with the highest values observed in stage IV patients. This stage-dependent pattern supports the hypothesis that uPAR reflects tumor aggressiveness and progression rather than early tumor initiation. In contrast, circulating uPA levels did not show significant differences either between patients and controls or across tumor stages, indicating a more limited role of uPA as a systemic biomarker in CRC.

Importantly, no significant associations were observed between circulating uPAR or uPA levels and key genomic tumor features, including tumor mutational burden (TMB), microsatellite instability (MSI), or homologous recombination deficiency (HRD). Furthermore, biomarker levels were not associated with mutation status of major CRC driver genes such as TP53, KRAS, BRAF, NRAS, PIK3CA, and FBXW7.

Together, these findings suggest that uPAR reflects biological processes that are largely independent of mutational burden and canonical genomic alterations. While TMB, MSI, and HRD primarily capture genomic instability and mutational processes, uPAR is more likely linked to tumor–microenvironment interactions, extracellular matrix remodeling, and invasive behavior.

The uPA/uPAR system is well known to regulate proteolytic activity, cell migration, and tissue remodeling, all of which are critical for tumor invasion and metastasis (71, 72). Elevated uPAR levels have been reported in multiple malignancies and are often associated with poor prognosis, increased metastatic potential, and reduced survival (73–75). Our results are consistent with these observations and further support the role of uPAR as a marker of tumor progression in CRC.

Interestingly, our previous studies in neuroblastoma models have revealed that uPAR/PLAUR expression is also associated with cancer stem cell properties and tumor dormancy, with low uPAR levels paradoxically correlating with an increased risk of recurrence (76, 77). This suggests that uPAR may play a dual role in tumor biology: high levels promote invasion and progression, whereas low levels may favour the persistence of quiescent, therapy-resistant stem-like cells that can drive late relapse. The interpretation of circulating uPAR levels in CRC should consider this complexity, as low uPAR might not simply indicate lower aggressiveness but rather a shift towards a dormant, stem- like state that predisposes to subsequent recurrence.

The lack of association between uPAR and genomic instability metrics, such as TMB and MSI, is particularly noteworthy. These results indicate that uPAR may provide information complementary to genomic biomarkers and could be especially valuable in patients with microsatellite-stable (MSS) tumors, where predictive biomarkers remain limited (18, 78, 79).

From a clinical perspective, the observed association between uPAR levels and tumor stage suggests its potential utility as a prognostic biomarker. Measurement of circulating uPAR could contribute to risk stratification, monitoring of disease progression, and potentially early detection of aggressive tumor behavior.

In addition, the uPA/uPAR axis represents a promising therapeutic target. Several experimental approaches aimed at inhibiting uPAR signaling or disrupting its interaction with ligands and co-receptors have demonstrated antitumor effects in preclinical models (80–83). Given its involvement in invasion and metastasis, targeting uPAR may complement existing therapies, particularly in advanced CRC.

Taken together, our findings highlight the importance of integrating molecular and biochemical markers to achieve a more comprehensive understanding of tumor biology. While genomic profiling provides insights into mutational processes, circulating biomarkers such as uPAR capture distinct aspects of tumor behavior related to invasion and microenvironmental interactions.

Further studies with larger cohorts and longitudinal sampling are needed to validate the prognostic value of uPAR and to explore its potential role in guiding therapeutic decision-making in colorectal cancer.

## Conclusion

This study demonstrates that circulating uPAR is a robust, stage-dependent biomarker of colorectal cancer progression that operates independently of canonical genomic features such as tumor mutational burden (TMB), microsatellite instability (MSI), homologous recombination deficiency (HRD), and major driver gene mutations (TP53, KRAS, BRAF, NRAS, PIK3CA, FBXW7). Unlike genomic markers that reflect mutational processes and genomic instability, uPAR likely captures tumor-microenvironment interactions, extracellular matrix remodeling, and invasive behavior, providing complementary information particularly valuable for microsatellite-stable tumors where predictive biomarkers are limited. Clinically, circulating uPAR shows potential as a prognostic biomarker for risk stratification, disease monitoring, and early detection of aggressive tumor behavior, while the uPA/uPAR axis represents a promising therapeutic target for inhibiting invasion and metastasis in advanced CRC. Future studies with larger cohorts and longitudinal sampling are needed to validate uPA prognostic value and explore its utility in guiding therapeutic decisions.

## Data Availability

All data produced in the present study are available upon reasonable request to the authors

## Author Contributions

PED - Data Curation, Formal Analysis, Writing – Original Draft; RKA - Formal Analysis, Writing – Original Draft; KVV - Data Curation; AMA - Data Curation; KPS - Data Curation, Formal Analysis; SVYu - Data Curation; KA - Data Curation; SEV – Conceptualization, Data Curation, Formal Analysis, Review & Editing. All authors have read and approved the final version of the manuscript and agree to be accountable for all aspects of the work.

## Funding

Research was supported from the Russian Federal Academic Leadership Program Priority 2030 at the Immanuel Kant Baltic Federal University and by Russian Federation under the State Assignment No. 03r-23/110-03 of Lomonosov Moscow State University.

## Acknowledgements

Language editing and translation assistance were provided using ChatGPT-4 (OpenAI) to improve grammar, clarity, readability, and overall language quality. All content was subsequently reviewed, revised, and approved by the authors, who take full responsibility for the accuracy, integrity, and final content of the manuscript.

## Conflicts of Interest

The authors declare no conflict of interest.

## Notes

### Competing Interest Statement

The authors have declared no competing interest.

### Author Declarations

This study had a cross-sectional design and was conducted in the Kaliningrad region within the framework of the National Genetic Initiative (NGI) population genomic program. Patients were recruited as part of the prospective observational study Factors Affecting the Results of Treatment of Patients With Colorectal Cancer (ClinicalTrials.gov identifier: NCT06050447). The study was conducted at Immanuel Kant Baltic Federal University in Kaliningrad in 2023. The study protocol was approved by the Local Ethics Committee of Immanuel Kant Baltic Federal University (Protocol No. 43, dated October 18, 2023). All participants provided written informed consent for the use of biological material and genetic data for research purposes

